# Estimating the transmission dynamics of Omicron in Beijing, November to December 2022

**DOI:** 10.1101/2022.12.15.22283522

**Authors:** Kathy Leung, Eric H. Y. Lau, Carlos K. H. Wong, Gabriel M. Leung, Joseph T. Wu

**Author notes:** Correspondence: Kathy Leung.

## Abstract

We tracked the effective reproduction number *R*_*t*_ of SARS-CoV-2 Omicron BF.7 in Beijing in November – December 2022 by fitting a transmission dynamic model parameterized with real-time mobility data to (i) the daily number of new symptomatic cases on November 1-11 (when the zero-covid interventions were still strictly enforced) and (ii) the proportion of individuals who participated in online polls on December 10-22 and self-reported to have been previously test-positive since November 1. After the announcement of “20 measures”, we estimated that *R*_*t*_ increased to 3.44 (95% CrI: 2.82 – 4.14) on November 18 and the infection incidence peaked on December 11. The cumulative infection attack rate (i.e. the proportion of population who have been infected since November 1) was 43.1% (95% CrI: 25.6 – 60.9) on December 14 and 75.7% (95% CrI: 60.7 – 84.4) on December 22. Surveillance programmes should be rapidly set up to monitor the evolving epidemiology and evolution of SARS-CoV-2 across China.

## Main Text

After implementing the “zero-COVID” strategy for more than two years, China has recently begun to adjust its COVID-19 response strategies, notably by announcing the “20 measures” on November 11 and further the “10 measures” on December 7, 2022 ^1,2^. Since then, Omicron infections spread rapidly in major cities of China, including Beijing, where the Omicron BF.7 epidemic has been putting great pressure on the healthcare system since early December ^3,4^.

Regular mass testing and intensive contact tracing were suspended in late November and nucleic acid testing has been conducted on a voluntary basis thereafter ^5^. As such, the daily number of confirmed cases reported by Beijing Municipal Health Commission (BMHC, http://wjw.beijing.gov.cn/) was no longer an accurate reflection of the epidemic, making it difficult to assess the transmission dynamics. Here we tracked the effective reproduction number of Omicron BF.7 in Beijing in November – December 2022 using our previous epidemic nowcast framework which combined real-time mobility data and case data in the disease transmission models ^6^.

### The effective reproduction number *R*_*t*_ of Omicron in Beijing

We categorized the COVID-19 response adjustments in Beijing in three stages:

i. **Stage 1** (November 1-11): Zero-COVID strategy was strictly implemented with regular mass testing, intensive contact tracing and lockdown of residential buildings or communities once PCR-positive infections and their close contacts were traced.
ii. **Stage 2** (November 12-25): Although mass testing and contact tracing were maintained after the announcement of “20 measures” on November 11, lockdown was limited to the residential buildings or only the floors or units where PCR-positive infections lived.
iii. **Stage 3** (after November 25): The requirement of regular mass testing, intensive contact tracing and lockdown were gradually relaxed and finally suspended on November 30. The nucleic acid testing has been conducted only on a voluntary basis after the announcement of “10 measures” on December 7.

We parameterised the disease transmission model with daily number of passengers from Beijing MTR (https://www.mtr.bj.cn/) and Beijing Subway (https://www.bjsubway.com/), which manage 22 of 27 subway lines in Beijing (**Figure 1** and **Supplementary Table 2**). We fitted the model to two data streams to estimate the effective reproduction number *R*_*t*_: i) the daily number of symptomatic cases reported to BMHC between November 1 and 11 (stage 1) when the testing and reporting behaviour were relatively stable; and ii) the proportion of participants who self-reported to be ever positive by polymerase chain reaction (PCR) or rapid antigen test (RAT) since November 1, based on Weibo and WeChat online polls conducted via convenience sampling between December 10 and 22 (**Figure 1**). See **Methods** for details.

**Figure 1.**
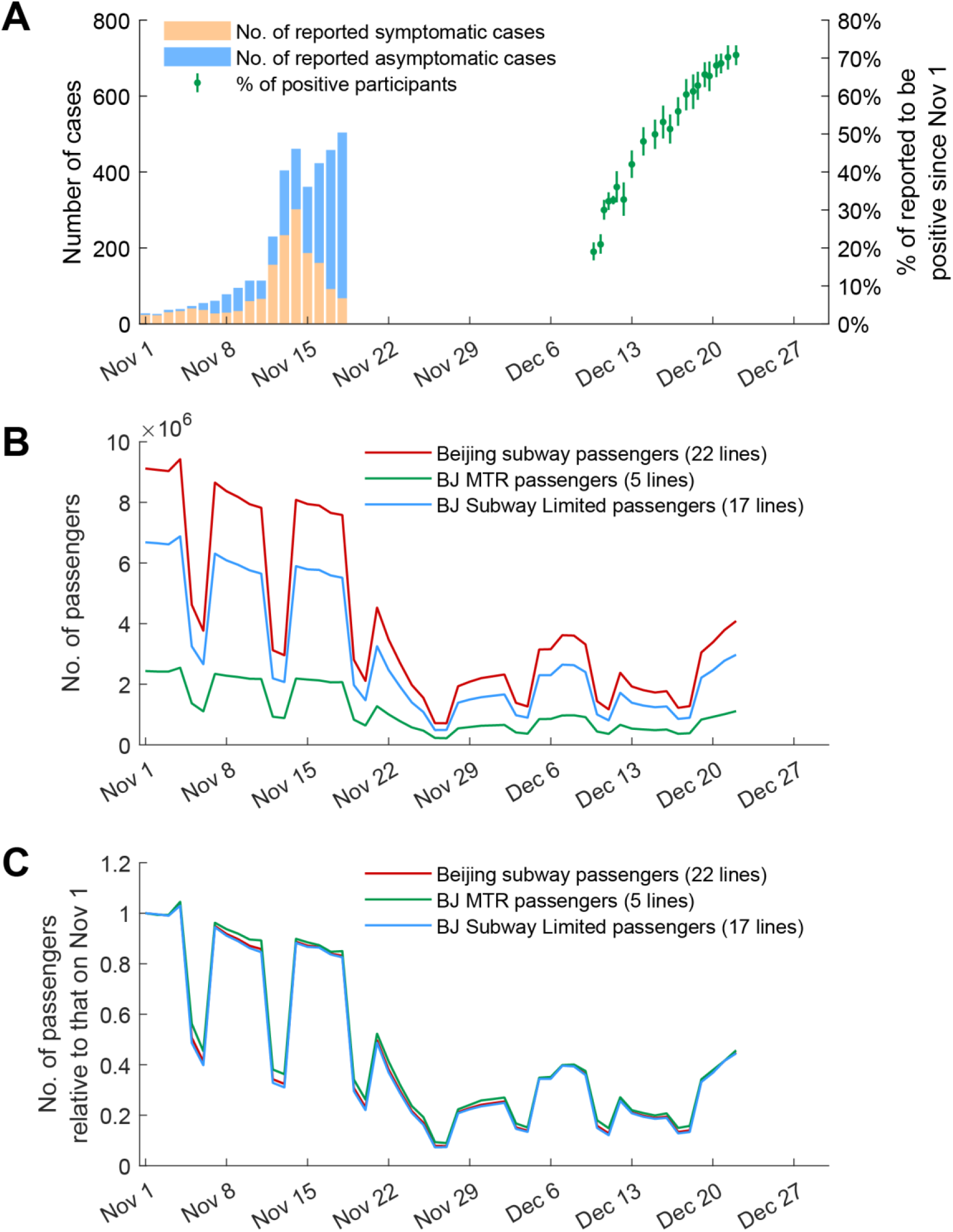
Data used in the inference. **(A)** The daily number of reported symptomatic cases between November 1 and 18 (orange), the daily number of reported asymptomatic cases between November 1 and 18 (blue), and the proportion of participants of Weibo or WeChat polls who reported to be ever tested positive (PCR or RAT) between December 10 and 22 (green). The details of Weibo or WeChat polls were provided in **Supplementary Table 1**. Bars in green indicate the 95% confidence interval (CI) of the proportions. **(B)** The daily number of passengers of Beijing subway lines managed by Beijing MTR (5 lines, https://www.mtr.bj.cn/) and Beijing Subway (17 lines, https://www.bjsubway.com/). **(C)** The daily number of subway passengers relative to that on November 1. The daily number of passengers were provided in **Supplementary Table 2**.

Within one week after the announcement of “20 measures”, *R*_*t*_ increased from 1.04 (95% CrI: 0.84 – 1.29) on November 11 to 3.44 (95% CrI: 2.82 – 4.14) on November 18 (**Figure 2**). In response to the increasing number of cases, public health and social measures (PHSMs) were implemented: residents were urged to stay home over the weekend of November 19-20; 95% of staff were advised to work from home during the week of November 21-25; kindergartens, primary and secondary schools were closed on November 21. Consequently, mobility decreased and *R*_*t*_ dropped to 0.99 (95% CrI: 0.83 – 1.16) on November 27.

**Figure 2.**
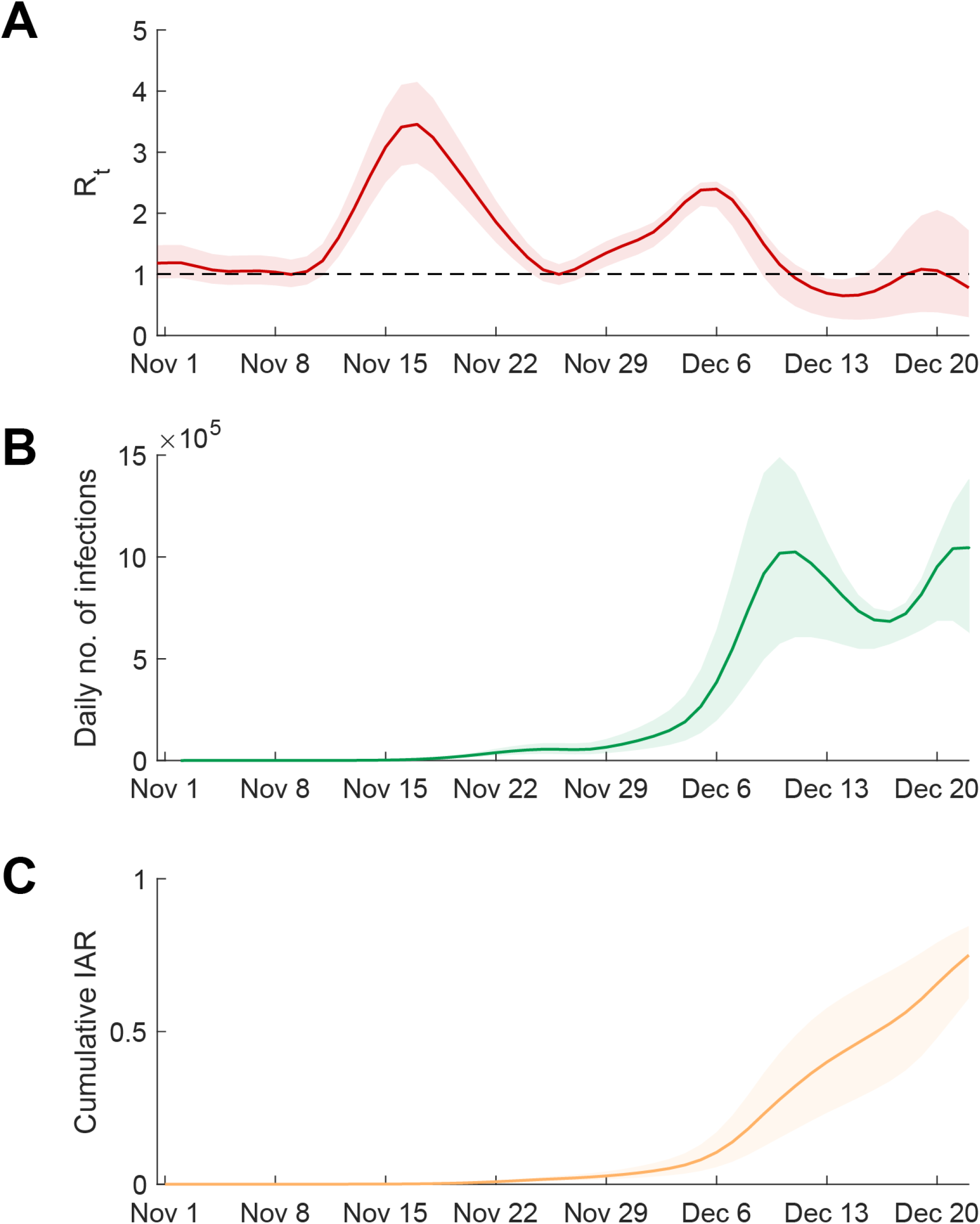
Estimated effective reproduction number *R*_*t*_, daily incidence and cumulative infection attack rate in Beijing. **(A)** Estimated *R*_*t*_ between November 1 and December 22. **(B)** Estimated daily number of infections between November 1 and December 22. **(C)** Estimated cumulative infection attack rates between November 1 and December 22. Lines indicate the *maximum a posteriori probability (MAP)* estimates and shades indicate the 95% credible interval (95% CrI).

The surge of infections saturated the capacity of PCR testing and quarantine facility in late November. The requirement of regular mass testing, intensive contact tracing and lockdown were gradually relaxed and finally suspended with the announcement of “10 measures”. PHSMs were relaxed and consequently *R*_*t*_ increased to 2.43 (95% CrI: 2.11 – 2.52) on December 7 (**Figure 2**), which was substantially higher than *R*_*t*_ of 1.9 under similar PHSMs in the early stage of Hong Kong’s Omicron wave in February – March 2022 ^7^.

Omicron infections spread rapidly again starting from early December, and many symptomatic individuals and their close contacts self-isolated. Within one week after the announcement of “10 measures”, Beijing MTR’s mobility dropped to low levels, and we estimated that *R*_*t*_ dropped below 1 on December 11 and to 0.72 (95% CrI: 0.30 – 0.93) on December 14. The mobility level started to increase in the week of December 17-23, and *R*_*t*_ increased to 1.04 (95% CrI: 0.39 – 1.96) on December 20.

### The daily incidence and cumulative infection attack rate

We estimated the daily incidence and cumulative infection attack rate from the fitted model accordingly (**Figure 2**). On November 30, when regular mass testing was suspended, we estimated the daily number of infections was 94,272 (95% CrI: 52,270 – 160,042). The daily incidence increased rapidly since then and peaked on December 11, with an estimated daily number of infections of 1.03 million (95% CrI: 0.61 – 1.49), i.e. 4.7% of the population.

We estimated that the cumulative infection attack rate (IAR) was 43.1% (95% CrI: 25.6 – 60.9) on December 14. The daily incidence slightly decreased between December 11 and 16 but started to increase again on December 17 due to increased mobility. We estimated the cumulative IAR has reached 75.7% (95% CrI: 60.7 – 84.4) on December 22.

In the base case scenario above, we assumed each participant of the online polls underwent multiple RATs and the ascertainment probability of previous infection was 1 after November 11 (**Figure 2**). As such, the actual IAR was likely to be higher than our base case estimates. As a sensitivity analysis, we included the ascertainment probability in the inference framework (**Supplementary Figure 1**). The resulting estimate of ascertainment probability was 81.5% (95% CrI: 78.9 – 84.2). The corresponding estimated *R*_*t*_ was slightly higher, e.g. 3.92 (95% CrI: 3.19 - 4.73) vs. 3.44 (95% CrI: 2.82 – 4.14) on November 18, and 3.05 (95% CrI: 2.63 – 3.24) vs. 2.43 (95% CrI: 2.11 – 2.52) on December 7. The estimated cumulative IAR was higher accordingly, reaching 55.1% (95% CrI: 34.3 – 73.5) on December 14 and 88.5% (95% CrI: 80.1 – 93.0) on December 22.

## Discussion

Our study tracked *R*_*t*_ of the Omicron BF.7 epidemic in Beijing by fitting an epidemic transmission model parameterised with mobility data to early-stage case count and recent survey data on cumulative incidence ^6^. When mass testing and confirmation of all cases become impossible during the surge of infections, it is important to continuously monitor infection prevalence in the community through various surveillance programmes, e.g. REACT-type studies in the UK and Hong Kong ^8,9^, wastewater surveillance ^10^, and serological surveillance ^11^. Given the roll-out of the second booster vaccines, data from such surveillance studies could also inform the assessment of vaccine effectiveness in real time ^9,11^.

By December 22, we estimated that the Omicron epidemic had peaked in Beijing and 76% of the Beijing residents had been infected (**Figure 2**). Assuming no changes in PHSMs nor population behaviour, we anticipated that the number of infections would start dropping towards the end of December and the pressure on the healthcare system would be alleviated. However, vaccination should be ramped up in the coming months in anticipation of upcoming waves that might arise due to importation of different Omicron variants from increased population movement between Beijing and other provinces during the Spring Festival in January 2023.

Our study has several limitations. First, passenger statistics of Beijing MTR and Beijing Subway were the only real-time mobility data we found. But these passengers only account for about 95% of Beijing’s subway volume and might not be representative of the population using other transportation means. Second, the subway mobility data might not be the best proxy for changes in contacts and hence *R*_*t*_. During the week of December 8-14, most of population stayed home or self-quarantined, and only made contact with their household members. Thus it is difficult to accurately estimate the rapid changes in population mixing and PHSM effectiveness. Third, our estimation relied on IARs inferred from the results of Weibo and WeChat online polls. Since most active Weibo or WeChat users are between 18 and 60 years old, the poll results might not be representative of all age groups. Self-reported infection status or test results can also introduce bias, and the ascertainment probability of infection was uncertain. Future refinements of our framework should include more reliable data to estimate IARs, such as age-specific seroprevalence data ^11^.

Given that COVID-19 response strategies have been adjusted nationwide, the number of new infections are expected to surge across China. The epidemics in other major cities and counties might be similar to that of Beijing with multiple incidence peaks, especially when population mixing starts to increase after the first peak which is further exacerbated by the upcoming mass population movement during the Spring Festival. Surveillance programmes should be rapidly set up to monitor the spread and evolution of SARS-CoV-2 infections, and further work should be done to track the transmissibility, incidence and IAR of the epidemics.

## Methods

### The transmission model

We used our previous age-structured SIR model to simulate the transmission of Omicron in Beijing ^12^:

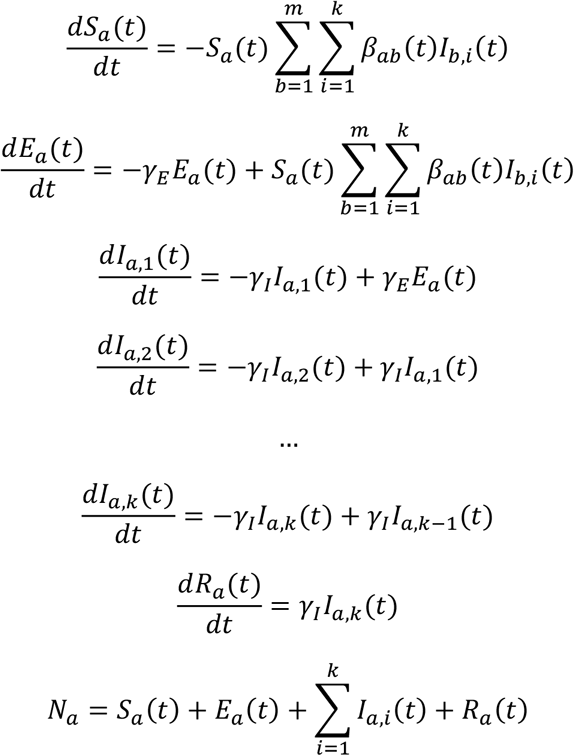

- *m* was the number of age groups in the population.
- *k* = 4 was the number of infectious states of the model.
- *S*_*a*_(*t*), *E*_*a*_(*t*) and *R*_*a*_(*t*) were the number of susceptible, exposed and removed individuals in age group *a* at time *t*.
- *I*_*a,i*_ (*t*) was the number of individuals in the *i*-th infectious state of age group *a* at time *t*.
- *N*_*a*_ was the total number of people in age group *a*.
- 1/γ_*E*_ was the duration from exposure to become infectious.
- *k*/γ_*I*_ was the duration of being infectious.
- The mean generation time 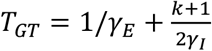, following the formulation of Svensson ^13^.

Following our previous framework to parameterise SARS-CoV-2 transmission models with mobility data ^6^, we formulated the average rate at which an individual in age group *a* made infectious contacts with age group *b* at time *t*, i.e., *β*_*ab*_(*t*) as

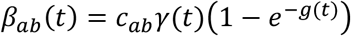

where

- *c*_*ab*_ was the contact rates between age group *a* and age group *b* from Mistry et al ^14^.
- *g*(*t*) was the normalized number of passengers of subway lines managed by Beijing MTR on day *t* (such that *g*(*t*) = 1 on November 1, 2022).
- γ(*t*) was the scaling factor for the mobility data proxy *g*(*t*). Given the changes in PHSMs during the weeks of November 12-25, we assumed γ(*t*) was γ_1_ between November 1 and 11, increased linearly between November 12 and 25 from γ_1_ to γ_2_, and remained at γ_2_ after November 25. γ_1_ and γ_2_ were estimated in the parameter inference **(Supplementary Table 3)**.

The time-varying next generation matrix for this SIR model was:

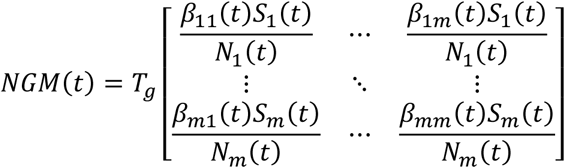

where *T*_*g*_ was the mean generation time.

The effective reproduction number *R*_*t*_ corresponded to the dominant eigenvalue of *NGM*(*t*) ^15,16^. The incidence rate of infection and reported onsets at time *t* were calculated as follows:

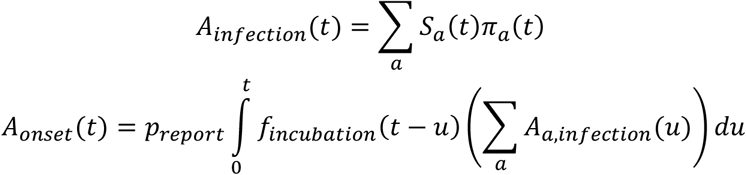

where *p*_*report*_ was the proportion of infections ascertained and reported as symptomatic cases by Beijing Municipal Health Commission (http://wjw.beijing.gov.cn/). Similarly, the cumulative infection attack rate at time *t* was calculated as follows:

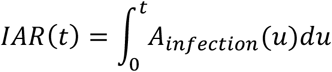

We assumed that the epidemic was seeded by *M* local infections on November 1, 2022.

### The inference

The set of parameters that were subject to statistical inference, which we denoted by *θ*, included: (i) the seed size *M*; (ii) the scaling factors γ_1_ and γ_2_; and (iii) the ascertainment proportion *p*_*report*_ between November 1 and 11 **(Supplementary Table 3)**. We estimated *θ* from (i) the daily number of symptomatic cases reported to Beijing Municipal Health Commission between November 1 and 11 **(Figure 1)** and (ii) the proportion of participants who reported to be positive by PCR or RAT since November 1 in Weibo or WeChat online polls conducted between December 10 and 22 **(Supplementary Table 1)**.

The likelihood function *L*(*θ*) is a product of the two components *L*_1_(*θ*) and *L*_2_(*θ*). *L*_1_(*θ*) was formulated as below:

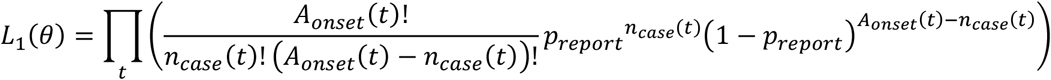

where

- *n*_*case*_(*t*) was the daily number of symptomatic cases confirmed and reported by Beijing Municipal Health Commission between November 1 and 11.
- *A*_*onset*_(*t*) was daily number of infections from the model convoluted by the incubation period distribution, assuming no onset-to-confirmation delay under mass testing and intensive contact tracing between November 1 and 11.

Similarly, assuming each participant of the online polls underwent multiple RATs and the ascertainment probability of infection was 100% after November 11, *L*_2_(*θ*) was formulated as below:

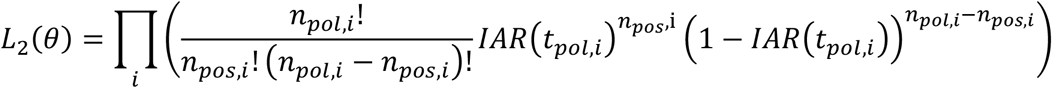

where

- *t*_*pol,i*_ was the time or date when Weibo or WeChat online poll *i* was carried out.
- *n*_*pol,i*_ was the number of participants of Weibo or WeChat online poll *i*.
- *n*_*pos,i*_ was the number of participants of Weibo or WeChat online poll *i* who reported to be ever tested positive by PCR or RAT since November 1.

However, the sensitivity of RAT ranges between 70% and 80% for Omicron and thus the ascertainment probability of infection could be less than 100% after November 11 ^17^. In a sensitivity analysis, we assumed that the ascertainment probability of infection (*pascertain*) was not 100%, and estimated *pascertain* and other parameters with the updated *L*_2_(*θ*) as below accordingly:

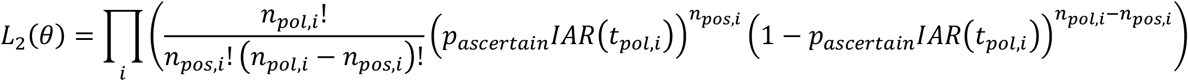

And *L*(*θ*) was formulated as below:

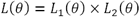

## Data Availability

We collated all data from publicly available data sources. All data included in the analyses are available in the main text or the supplementary materials.

## Contributors

All authors designed the study, developed the model, analysed data, interpreted the results, and wrote the manuscript.

## Data sharing statement

We collated all data from publicly available data sources. All data included in the analyses are available in the main text or the supplementary materials. All data and MATLAB codes are available at https://github.com/kathyleung/2022_12_24_Beijing_Rt_Omicron.

## Supplementary Material

**Supplementary Table 1.** The daily proportion of participants of Weibo or WeChat online polls who reported to be tested positive by PCR or RAT since November 1, 2022, in Beijing.

| Date | Proportion of participants who reported to be positive | No. of participants who reported to be positive | No. of participants | Source and Weibo online polls |
| --- | --- | --- | --- | --- |
| Dec 10 | 0.190 | 211 | 1110 | https://weibo.com/u/2987102112 |
| Dec 10 | 0.210 | 214 | 1021 | https://weibo.com/u/7126731879 |
| Dec 11 | 0.300 | 373 | 1243 | https://weibo.com/u/2987102112 |
| Dec 11 | 0.320 | 524 | 1622 | https://weibo.com/chinaetfs |
| Dec 11 | 0.330 | 2296 | 7043 | https://weibo.com/chinaetfs |
| Dec 12 | 0.360 | 199 | 552 | https://weibo.com/u/2987102112 |
| Dec 12 | 0.320 | 150 | 458 | https://weibo.com/u/7126731879 |
| Dec 13 | 0.420 | 311 | 740 | https://weibo.com/u/2987102112 |
| Dec 14 | 0.480 | 345 | 718 | https://weibo.com/u/2987102112 |
| Dec 15 | 0.499 | 331 | 663 | https://weibo.com/u/2987102112 |
| Dec 16 | 0.531 | 285 | 536 | https://weibo.com/u/2987102112 |
| Dec 16 | 0.513 | 348 | 678 | https://mp.weixin.qq.com/s/PJuDIocTon-WaWTRLldXMw |
| Dec 17 | 0.560 | 389 | 695 | https://weibo.com/u/2987102112 |
| Dec 18 | 0.605 | 342 | 566 | https://weibo.com/u/2987102112 |
| Dec 18 | 0.612 | 281 | 459 | https://mp.weixin.qq.com/s/PJuDIocTon-WaWTRLldXMw |
| Dec 19 | 0.628 | 430 | 685 | https://weibo.com/u/2987102112 |
| Dec 19 | 0.657 | 555 | 845 | https://mp.weixin.qq.com/s/PJuDIocTon-WaWTRLldXMw |
| Dec 20 | 0.653 | 391 | 599 | https://weibo.com/u/2987102112 |
| Dec 20 | 0.681 | 656 | 964 | https://mp.weixin.qq.com/s/PJuDIocTon-WaWTRLldXMw |
| Dec 21 | 0.686 | 857 | 1249 | https://weibo.com/u/2987102112 |
| Dec 21 | 0.703 | 583 | 830 | https://mp.weixin.qq.com/s/6jSGzWt_kyd6K880gnQ6wA |
| Dec 22 | 0.708 | 861 | 1216 | https://weibo.com/u/2987102112 |

**Supplementary Table 2.** The daily number of passengers of Beijing Subway including 5 lines managed by Beijing MTR Corporation Limited and 17 lines managed by Beijing Mass Transit Railway Operation Corporation Limited (‘0000) *.

| **Date** | **Passengers of 5 lines managed by Beijing MTR Corporation Limited** | **Passengers of 17 lines managed by Beijing Mass Transit Railway Operation Corporation Limited** |
| --- | --- | --- |
| 11/1/2022 | 243.5 | 668.14 |
| 11/2/2022 | 241.9 | 665.35 |
| 11/3/2022 | 241.8 | 661.44 |
| 11/4/2022 | 254.5 | 688.01 |
| 11/5/2022 | 137 | 325.2 |
| 11/6/2022 | 110.5 | 265.85 |
| 11/7/2022 | 234.3 | 631.04 |
| 11/8/2022 | 228.1 | 608.73 |
| 11/9/2022 | 223.8 | 594.07 |
| 11/10/2022 | 218.1 | 575.84 |
| 11/11/2022 | 217.3 | 564.99 |
| 11/12/2022 | 92.8 | 219.41 |
| 11/13/2022 | 88.3 | 207.25 |
| 11/14/2022 | 218.8 | 589.57 |
| 11/15/2022 | 215.4 | 579.04 |
| 11/16/2022 | 212.7 | 577.11 |
| 11/17/2022 | 206.3 | 558.94 |
| 11/18/2022 | 206.9 | 551.43 |
| 11/19/2022 | 83.1 | 197.43 |
| 11/20/2022 | 63.8 | 147.13 |
| 11/21/2022 | 127.4 | 325.61 |
| 11/22/2022 | 100.5 | 246.89 |
| 11/23/2022 | 77.8 | 191.41 |
| 11/24/2022 | 57.6 | 140.05 |
| 11/25/2022 | 46.9 | 108.3 |
| 11/26/2022 | 22.8 | 48.77 |
| 11/27/2022 | 21.9 | 49.33 |
| 11/28/2022 | 54.4 | 138.99 |
| 11/29/2022 | 58.7 | 149.52 |
| 11/30/2022 | 63 | 157.22 |
| 12/1/2022 | 64.3 | 161.86 |
| 12/2/2022 | 65.8 | 166.36 |
| 12/3/2022 | 40.8 | 97.72 |
| 12/4/2022 | 36.9 | 89.69 |
| 12/5/2022 | 84.9 | 229.71 |
| 12/6/2022 | 85.6 | 230 |
| 12/7/2022 | 97.1 | 264.6 |
| 12/8/2022 | 97.6 | 262.9 |
| 12/9/2022 | 91.4 | 239.95 |
| 12/10/2022 | 43.9 | 100.15 |
| 12/11/2022 | 36.2 | 80.83 |
| 12/12/2022 | 66.1 | 171.78 |
| 12/13/2022 | 53.6 | 138.92 |
| 12/14/2022 | 50.8 | 129.62 |
| 12/15/2022 | 48.5 | 124.21 |
| 12/16/2022 | 50.5 | 126.64 |
| 12/17/2022 | 36.5 | 85.66 |
| 12/18/2022 | 38.4 | 89.35 |
| 12/19/2022 | 83.3 | 221.57 |
| 12/20/2022 | 92.3 | 246.03 |
| 12/21/2022 | 101.2 | 277.56 |
| 12/22/2022 | 111.2 | 297.2 |

\* Data from https://weibo.com/u/2125939111, https://www.mtr.bj.cn/, https://weibo.com/u/2778292197 and https://www.bjsubway.com/.
The map of all of the 27 subway lines is available at:
http://www.bjmoa.cn/indishare/sitemaster.nsf/frmthiridxlt_xm?openform&database=bjmtroa/lxt.nsf&view=vwPublicedByCatforsite3_xm&path=1code&mkfl=ccfw

**Supplementary Table 3.** Parameters fitted in the transmission model.

| Parameter | Description | Estimate (95% CrI) |
| --- | --- | --- |
| $\gamma_1$ | The scaling factor for translating the mobility data proxy into the contact matrix between November 1 and 11 (see methods) | 0.0285<br>(0.0269 – 0.0304) |
| $\gamma_2$ | The scaling factor for translating the mobility data proxy into the contact matrix after November 25 (see methods) | 0.0951<br>(0.0942 – 0.0960) |
| $M$ | The number of local infections on November 1 that might generate community transmission when the simulation was started | 1562<br>(906 – 2292) |
| $p_{report}$ | The proportion of infections ascertained as symptomatic cases by Beijing Municipal Health Commission | 8.70%<br>(6.84 – 11.65) |

**Supplementary Table 4.** Fixed parameters in the transmission model.

| Parameter | Description, assumption, and source | Value |
| --- | --- | --- |
| $1/\gamma_E$ | Duration from exposure to becoming infectious | Assumed to be 1 day |
| $T_{GT}$ | Mean generation time <sup>8</sup> | 4.6 days<br>Assumed to be normal distributed with the coefficient of variation of 0.05 when the posterior distributions of $R_t$ and IARs were generated |
| $VE_S$ | Vaccine effectiveness in reducing susceptibility | Assumed to be 0 since most of the vaccinations were given >8 months ago and most of the vaccines were inactivated virus vaccines |
| $VE_I$ | Vaccine effectiveness in reducing infectiousness | Assumed to be 0 |
| $f_{incubation}$ | Probability density function of incubation period <sup>18,19</sup> | Gamma distribution<br>Mean: 3.5 days<br>SD: 2.6 days |

**Supplementary Figure 1.**
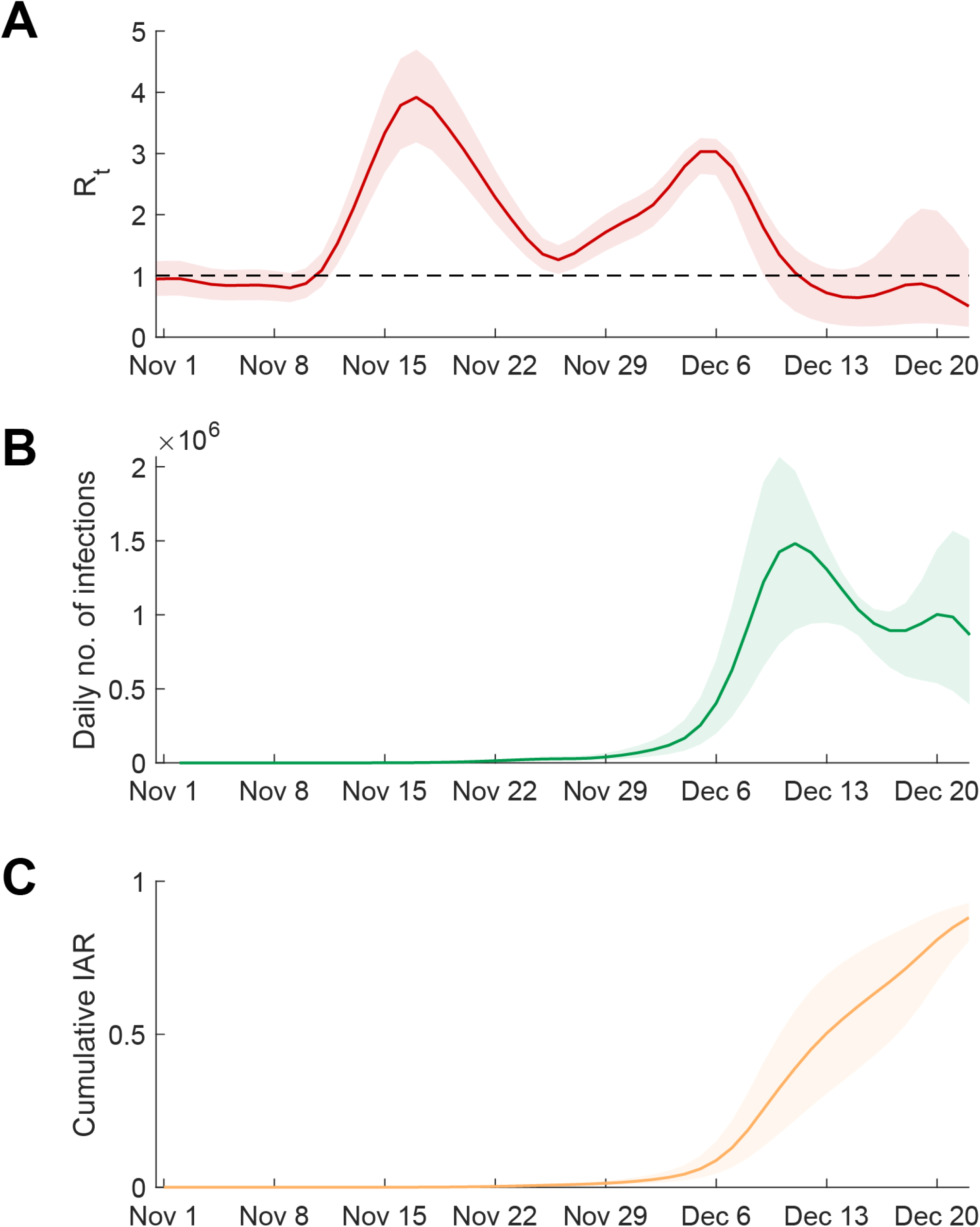
Estimated *R*_*t*_, daily incidence and cumulative infection attack rate in Beijing when the ascertainment probability after November 11 was estimated in the inference. **(A)** Estimated *R*_*t*_ between November 1 and December 22. **(B)** Estimated daily number of infections between November 1 and December 22. **(C)** Estimated cumulative infection attack rates between November 1 and December 22. Lines indicate the *maximum a posteriori probability (MAP)* estimates and shades indicate the 95% credible interval (95% CrI).

